# MODEL FAMILY CENTERED CARE IN CHILDREN WITH DIABETES MELITUS : A PHILOSOPHICAL PERSPECTIVE

**DOI:** 10.1101/2022.07.24.22277979

**Authors:** Siti Maimuna

**Affiliations:** Faculty of Nursing, Universitas Airlangga

**Keywords:** Nursing, family centered care, diabetes melitus, Children

## Abstract

**Introduction:** Diabetes mellitus is the most widespread chronic non-infectious disease, with an increase in the frequency in childhood that could be the epidemic of the 21st century. This literature aims to study the philosophy of family center care interventions to improve the quality of life of children with diabetes melitus viewed from three philosophical perspectives, namely ontology, epistemology, and axiology.

**Method:** A literature search was performed on five databases, namely SCOPUS, ProQuest, PubMed, ScienceDirect, SAGEPub, and Google Scholar. Population limitations and diagnoses in this literature of children with diabetes melitus. This research is a quantitative study focusing on publications between 2017-2022.

**Result:** Family centered care can improve the quality of life of children with diabetes. Children with diabetes successfully manage their disease is possible because parents have understood and received ongoing training and in this case, children with connections to the diabetes care team (family) and medical team play an important role in the management of children’s diabetes. Training and strengthening education helps families to control disease. Teaching children and their families to improve knowledge and control diabetes and metabolic diseases.

**Conclusion:** Family centered care for children with diabetes requires family knowledge about care, training skills, building strong motivation for children with diabetes so that complications do not occur.

## INTRODUCTION

Diabetes mellitus is the most widespread chronic non-infectious disease, with an increase in the frequency in childhood that could be the epidemic of the 21st century (Miolski et al., 2020). Each year an estimated 130,000 children and adolescents are newly diagnosed with Type 1 Diabetes (T1DM) globally (Palmer et al., 2022). Complications of diabetes include diabetic nephropathy, retinopathy, neuropathy, and peripheral arterial disease (PAD) (Mponponsuo et al., 2021). Diabetes is an incurable disease, but with adequate management and monitoring children can have a good quality of life. The goals of therapy in type 1 diabetes are to achieve optimal metabolic control, prevent acute complications, prevent long-term microvascular and macrovascular complications, and help psychologically for children and their families (Pulungan et al., 2019). In the area of pediatric nursing, using family-centered care (FCC) is a nursing philosophy based on the partnership between the family and the health care team in providing care for a sick child. Collaborative partnerships are based on dignity and respect, information sharing, and family participation through acquired competencies in providing care for sick children, including children with diabetes mellitus (Johnson & Abraham, 2012)

Even with adequate healthcare in place, T1DM is a challenging disease to manage. Maintaining glycemic control requires a variety of self-care strategies including exercise, meal plans, frequent blood glucose monitoring, and the storage, preparation, and administration of insulin. For children, meeting targets for glycemic control is also associated with high levels of parental support and involvement in disease management (Palmer et al., 2022). Parents often complain of distress, frustration, and feeling strange caring for a sick child with diabetes mellitus. However, if they are given the opportunity to be involved in care, receive clear communication about their child’s status from the health care team, and establish good relationships with health workers, they experience satisfaction and reduce stress. Implementation of the FCC has been shown to reduce length of hospital stay for sick children, improve maternal and child well-being, enable better allocation of human resources, and enhance parent-child bonding(Qian et al., 2021)

Philosophy is an attitude or view of life and an applied field to help individuals to evaluate their existence in a more satisfying way. Philosophy leads to understanding and understanding leads to appropriate action. In general, philosophical systematics has three main discussions or sections, namely; epistemology or theory of knowledge which discusses how to acquire knowledge, ontology or theory of nature which discusses the nature of everything that gives birth to knowledge and axiology or theory of value which discusses the use of knowledge. Thus, studying these three branches is very important in understanding such a wide scope of philosophy.(Babbie, 2020)

## METHOD

The research method used is an integrative literature review. This review proposes a family centered care intervention in children with diabetes mellitus. We systematically searched Scopus, PubMed, ProQuest, and Science Direct. Searches using various combinations of keywords with the help of Boolean operators, including: “Nursing” AND “family centered care” AND “Children’s diabetes mellitus”, were combined as terms. The inclusion criteria applied in this study were peer-reviewed articles in English that discussed family centered care and diabetes mellitus in children. Articles published in the last five years (2017-2021). Research studies are conducted in various fields that specifically examine family centered care interventions in children with diabetes mellitus. This study is a quantitative study with a randomized controlled trial (RCT) and full text method. The first author conducted an initial database search and articles for review. We used the PRISMA Flowchart 2009 (Moher et al., 2010) to record the article review and inclusion process (see Figure 1). An initial search of the four databases yielded 423 results. After that, we collect all articles and remove duplicate articles. Sources were excluded based on title and abstract if they were not peer-reviewed research studies or related to family-centered care interventions in children with diabetes mellitus within the scope of nursing. The next step is to narrow the selection of articles based on the year of publication and the context of the study. After the remaining articles were assessed for significant QoL findings in pediatric patients with diabetes mellitus, 7 papers were selected for final inclusion (Figure 1).

**Figure 1.**
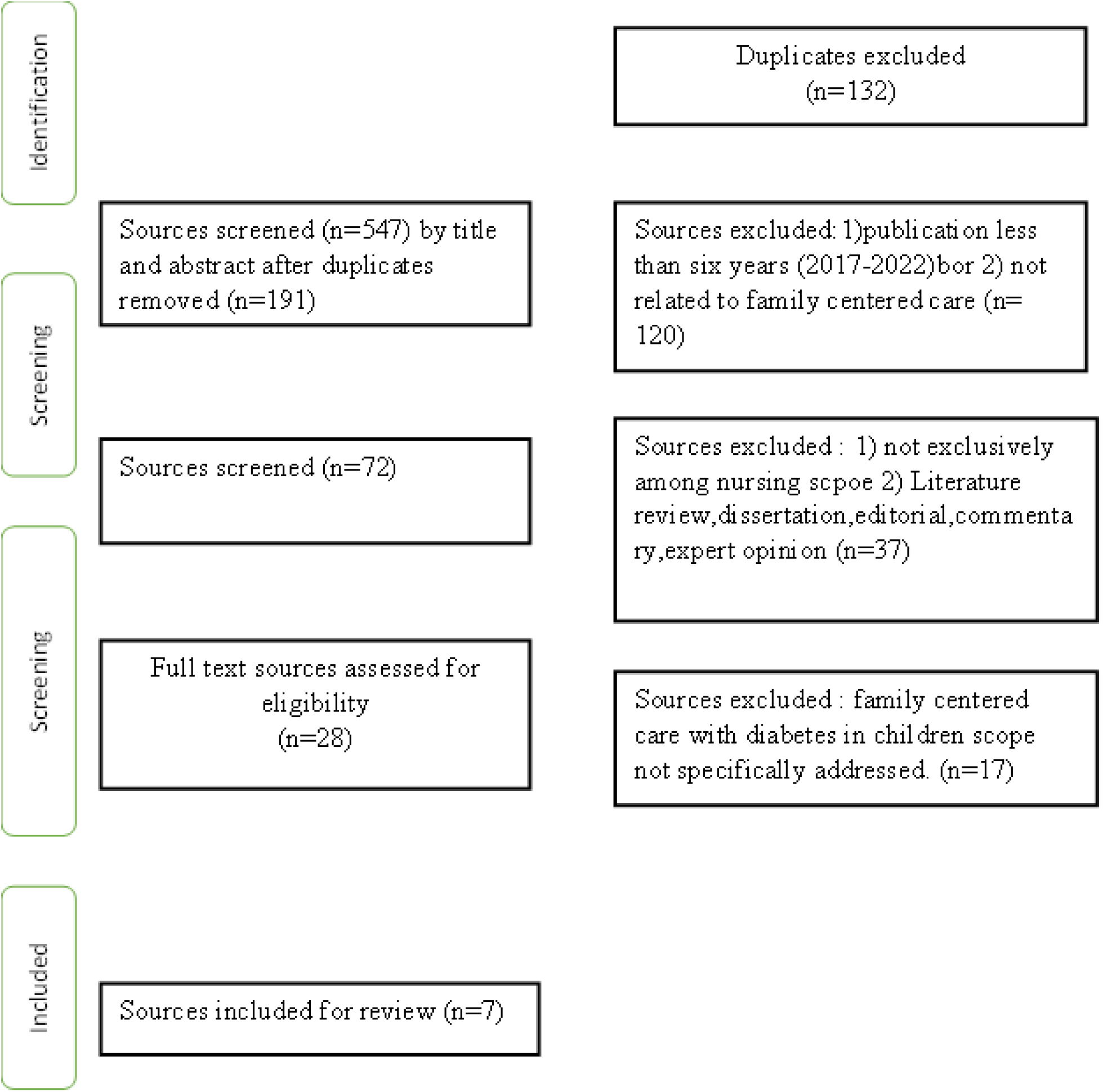
PRISMA Flowchart of Literature Search and Screening Process

**Figure 2.**
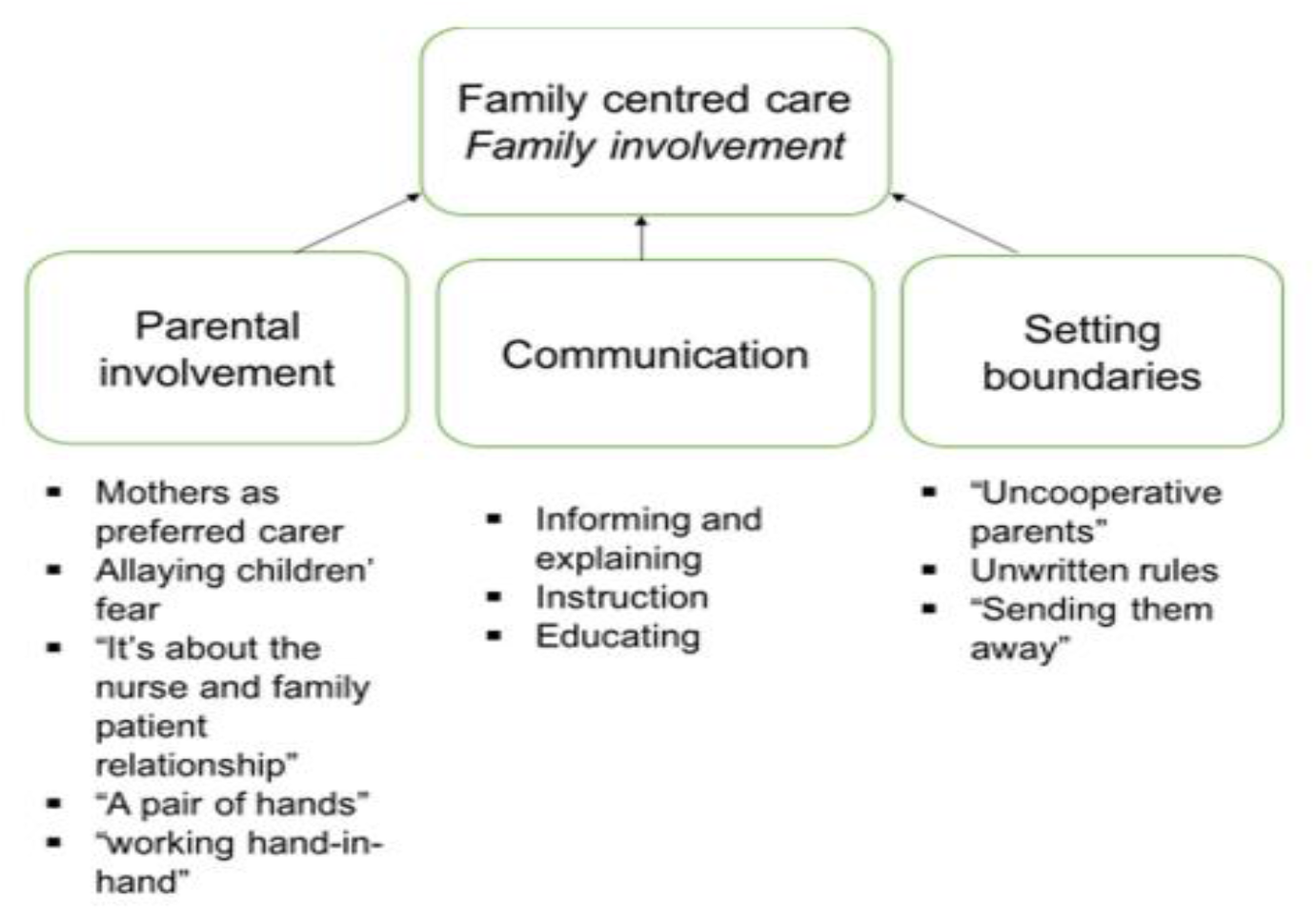
Family Centered Care (Ohene et al., 2020)

## RESULT AND DISCUSSION

### 1. Ontology Study Family Centered Care in Children with Diabetes Melitus

Family-centered care (FCC) is a philosophy of care based on a partnership between the family and the health care team in providing care to a sick child. This collaborative partnership is based on dignity and respect, information sharing, and family participation through their acquired competence in providing care for sick children with diabetes mellitus. (Sarin & Maria, 2019). One of the goals of the FCC is to build skills of parents and families in caring for and sick during inpatient care and to bring these skills into home care to maintain the continum of care. (Maria et al., 2021).

In the aspect of family centered care, there are two important concepts, namely the concept of enabling and empowering. The enabling concept views that the family has a role in the care provided. Nurses must involve the family in providing care in order to meet the needs of the child and the family in general. In the concept of empowering, nurses can involve the family in making decisions about the actions to be taken (Zimmerman, 2000). Family involvement is needed considering that children always need their parents. The FCC program establishes standardized care approaches and behavioral norms for health care providers that direct psychosocial and tangible support to parents so that a child is not separated from his or her parents against their will when receiving follow-up care. Introduction of family-centered care in the unit as a high priority to facilitate bonding and engagement with potential positive outcomes for both parent and child. (Maree et al., 2017)

The ethos of family centered care nursing care is basically care and giving parents a sense of security and comfort for their children is child nursing care so that nursing care must be centered on the concept of the child as part of the family and the family as the best support provider. Family Centered Care is defined as a philosophy of family-centered care, recognizing the family as a constant in a child’s life. Family Centered Care believes in individual support, respecting, encouraging and enhancing the strengths and competencies of families. Elements of family centered care include (1) incorporating an understanding into policy and practice that the family is a constant in the child’s life, (2) facilitating family/professional collaboration at all levels of nursing care, (3) exchanging complete and clear information between family members and caregivers professional. (Hei et al., 2021).

Family centered care can be applied in all age groups and all clinics. However, this model of care is very important in pediatric services because children depend on family members to meet their self-care and needs. Family-centered care is an approach that accepts family cultural differences and takes into account the needs of not only children, but all family members. It is a model of care that provides collaboration between parents and health professionals and an approach to children and their families as a physical, emotional, social, cultural and religious whole. Respect, cooperation and support form the basis of a family-centered care philosophy. (Kucuk Alemdar et al., 2018b)

Family nursing interventions are needed to overcome the problem of family powerlessness. This intervention is aimed at increasing the ability of families in the health sector, among others; the family’s ability to recognize the health problems they face, make the right decisions to deal with health problems and the ability to care for sick family members(Friedman et al., 2010). There is a positive effect of education based on the family centered empowerment model in the case group compared to conventional training methods. The role of the family in patient health, the application of this training method is recommended to nurses and authorities in order to facilitate optimal metabolic control in type 1 diabetes patients. Family empowerment interventions are based on the belief that every family has the potential and ability to develop and become more independent (Sargazi Shad et al., 2018)

### 2. Epistemological Study of Family Centered Care in Children with Diabetes Melitus

The practical application of the FCC requires the mutual dependence of the family and the health care system. Parents depend on the knowledge and expertise of professionals, and the latter depends on the emotional and physical attachment of parents to their children. Thus, collaboration or partnership is based on shared responsibility for the child. The FCC model was designed in a hospital where parents were provided with clear audio-visual information and repetitive interactive education, which had built their capacity to carry out care activities. (Sarin & Maria, 2019).

Nursing interventions using a family centered care approach emphasize that policy making, planning care programs, designing health facilities, and daily interactions between clients and health workers must involve the family. Families are given the authority to be involved in the care of clients, which means that families with family backgrounds, expertise and competencies provide positive benefits in child care. Giving authority to the family means opening the way for the family to know the strength, ability of the family in caring for children. (Fratantoni et al., 2022)

Philosophically, the involvement of parents and children in their own care is very important. This does not mean simply asking them what they think or talk about with parents and not for children (of course, this is age related). Partnerships between health professionals and children and parents are at the core of successful implementation of centered care. on the family. The onus is on the healthcare professional to put this in its place and make it work, because the power gradient of any healthcare interaction sees the healthcare professional on top providing care to the patient/client/family below. Therefore, family centered care will only be successful if health professionals can adapt all interactions according to the contingency and family situation, treatment, illness or condition and health services at the time. (Facn, 2015)

The family is the unit of society responsible for providing adequate health care for patients and their relatives. When providing care to a patient, the family needs a good understanding of the illness. Families who have a level of knowledge and education in learning and understanding the medical model in terms of providing a balanced diet, and monitoring drug therapy will be enhanced by being involved in providing nursing care, and creating a sense of usefulness as a team member. This approach will improve the health and well-being of the family. In accordance with the progress made in the field of health problems, caregivers in the family are a substitute for hospital care.(Hakim, 2017)

### 3. Axiologi Study of Family Centered Care in Children with Diabetes Melitus

**Table 1.**
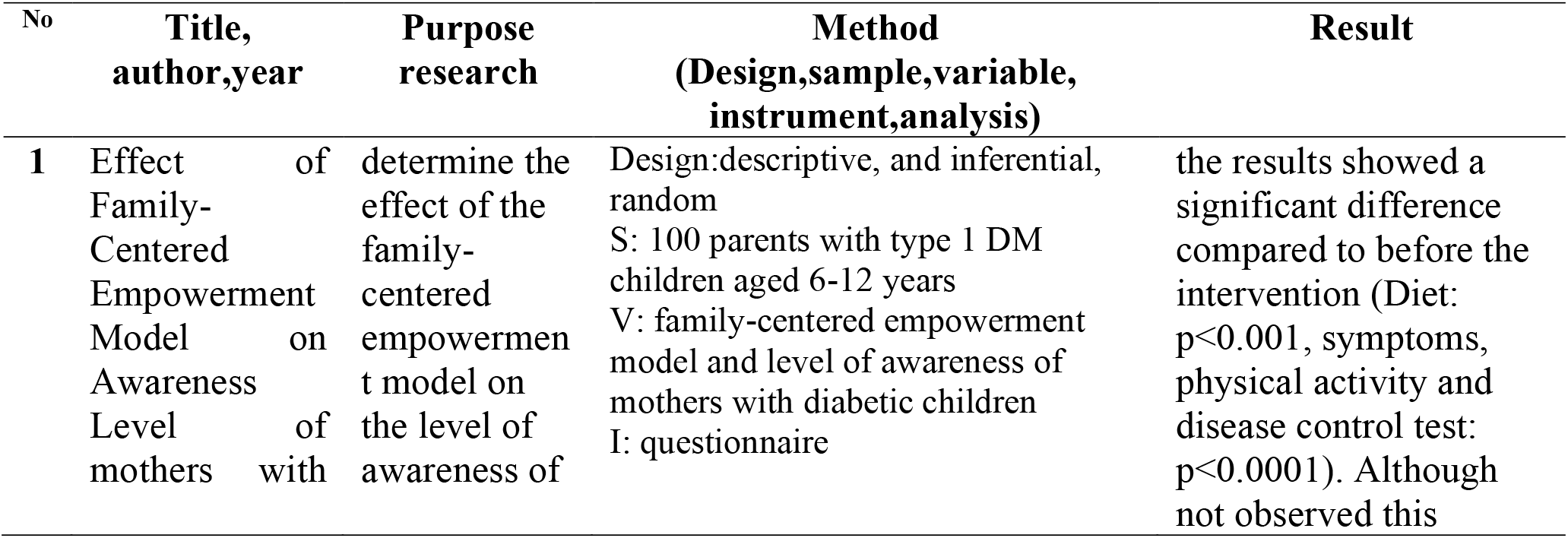
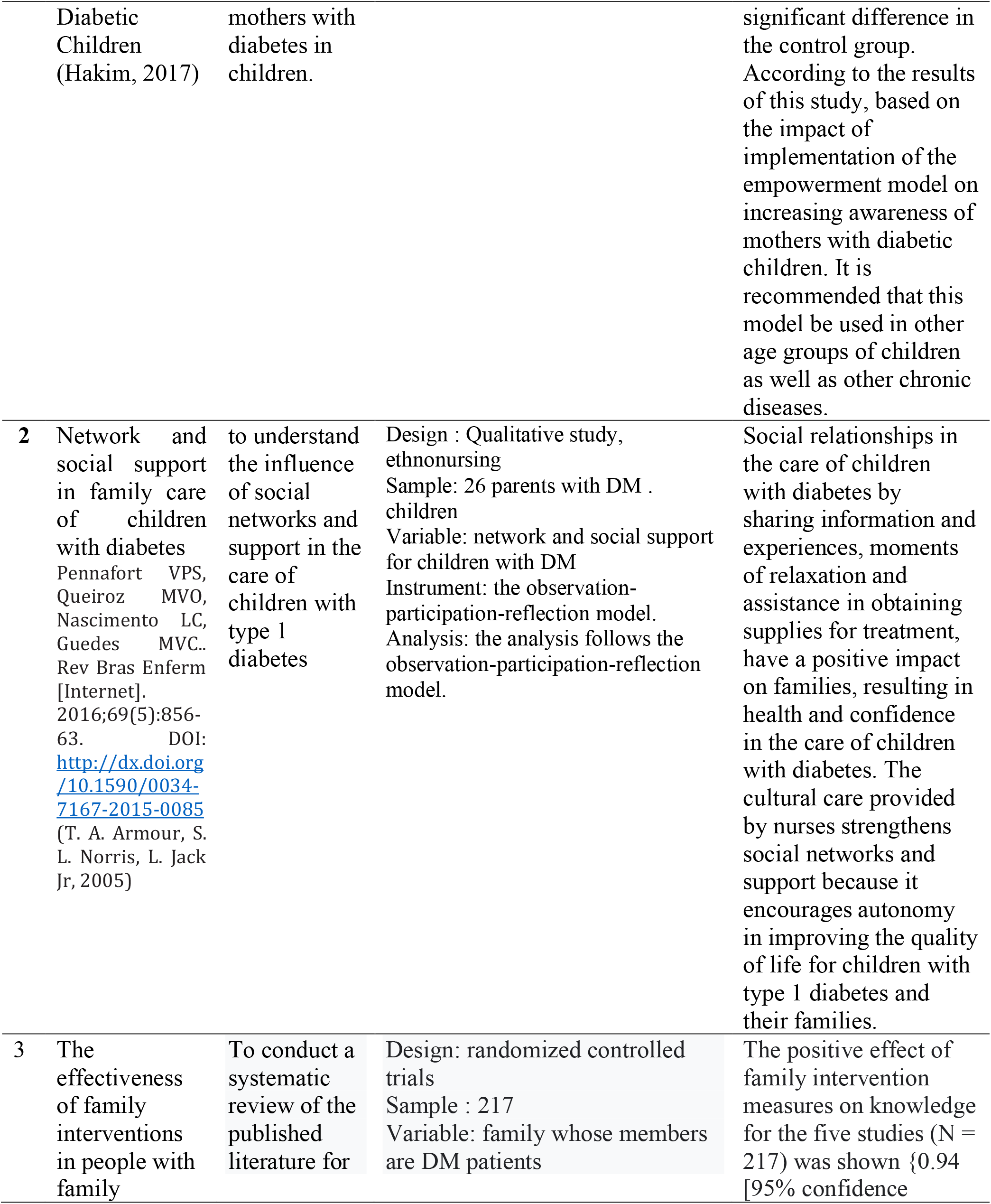
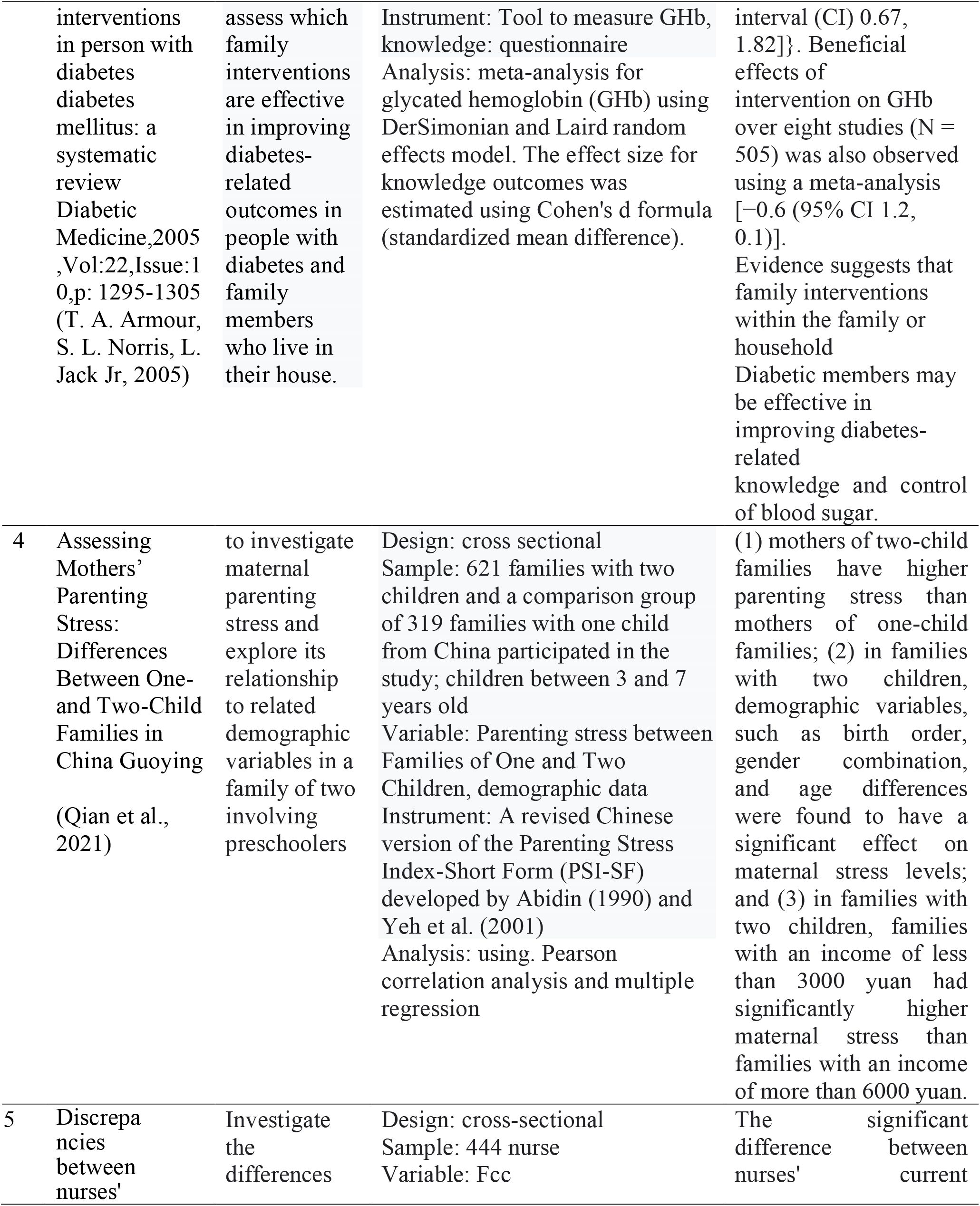
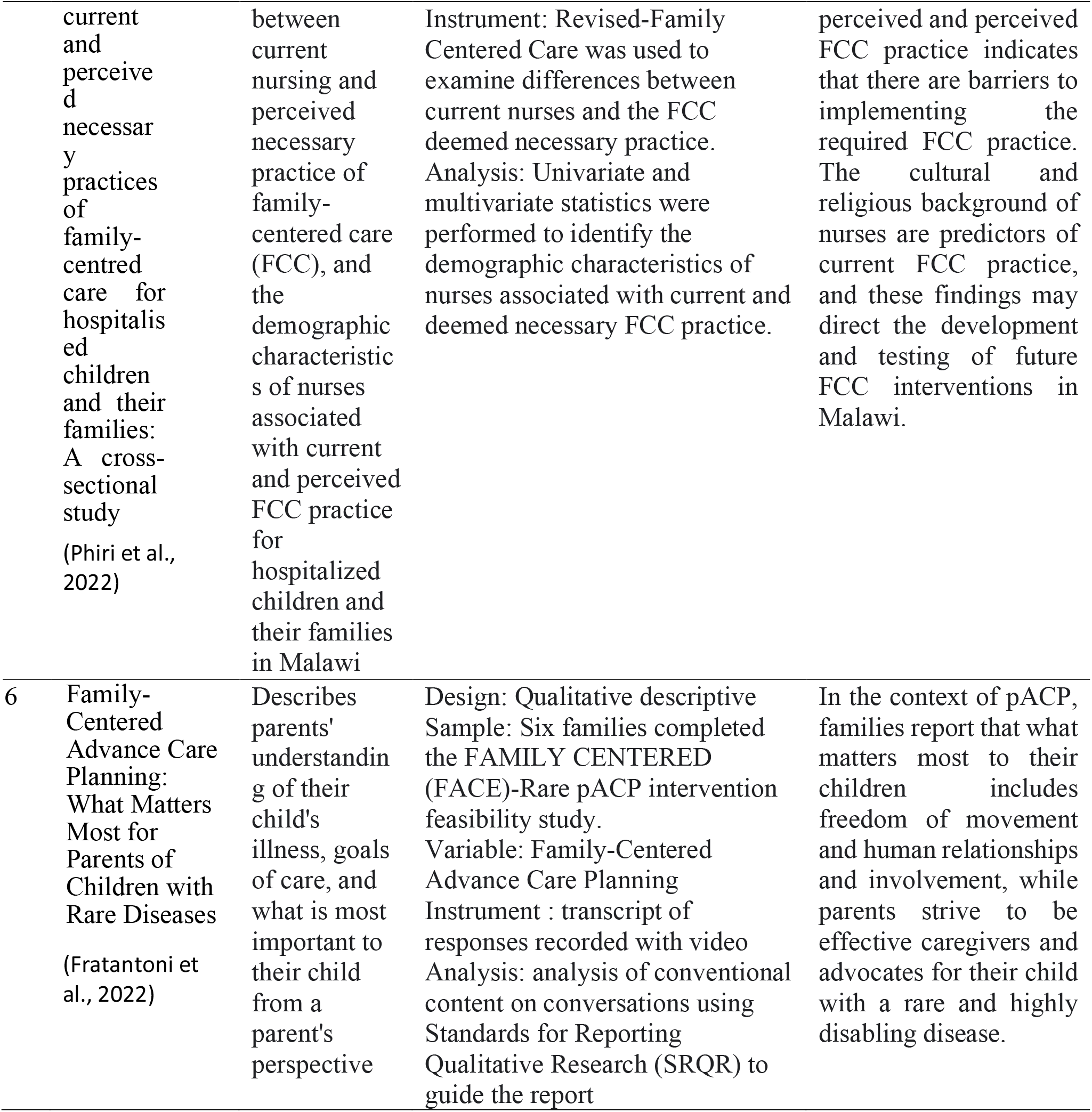
The results of the review of articles based on the Axiological perspective approach (n=6)

Parents often report distress, frustration, and alienation if they are excluded from caring for a sick child. However, if they are given the opportunity to be involved in care, receive clear communication about children suffering from diabetes mellitus from health professionals, and build relationships with health professionals, they experience satisfaction and reduce stress. FCC is beneficial for parents and nurses, namely reducing parental anxiety, assisting nurses in supervising children, and providing a positive response for children. In addition, the FCC has found that this approach results in increased performance satisfaction for nurses, although the direct impact of the intervention has been found to present challenges for nurses and health care providers, such as increased time away from patient care, difficulty in coordinating communication, and increased time to care. support and communicate with parents (Hei et al., 2021; Sarin & Maria, 2019).

Controlling diabetes means preventing and delaying its complications. Diabetes that is not well controlled can cause high blood sugar, which has a very strong relationship with chronic complications such as: cardiovascular complications, and retinopathy, nephropathy, neuropathy, and various long-term psychological effects that are complications associated with high health costs and poor quality of life. Changes due to complications can be delayed up to 20 years later with appropriate control, and if control is poor, vascular changes appear sooner than 2.5-3 years after diagnosis. It takes a great ability to control this disease so that it can be managed properly by involving the role of parents and children at home. The effectiveness of diabetes care, treatment and control depends on patient and family participation in the family center care model and family empowerment (Sargazi Shad et al., 2018)

The family empowerment model is based on three steps: threat perception, problem solving and evaluation. Perceived threat: perceived severity (i.e., perceived risk in the event of failure) of prevention is the first step in an empowerment model of treatment improvement. Lack of parental awareness of children with diabetes is one of the causes of their problems. This phase is carried out by increasing awareness of understanding and therefore understanding the severity through providing information on the nature of the disease, early and late complications of diabetes such as hypoglycemia, hyperglycemia, diabetic retinopathy, nephropathy, neuropathy and diabetic ketoacidosis and important issues about nutrition and exercise for children with diabetes (Miolski et al., 2020)

Generally, empowering the family system (patients and other family members) will improve their health status. Knowledge of mothers and their training has a significant impact on better diabetes and reducing the late complications of diabetes in children. Nurses as key figures in caring for patients help families, and increase hope and trust. Such an approach will improve the health and well-being of the family. Thus, nurses are in a unique position to interact with individual and family members. They can gain the awareness, skills and support needed to maintain the quality of care they provide at home; The goal of nursing interventions in family-centered care is to promote the ability of family members in specific areas to overcome existing barriers to health and well-being (Hakim, 2017)

An important role of nurses is the training of patients with diabetes and their families in various areas of disease and disease control measures. Children with diabetes successfully manage their disease is possible because parents have understood and received ongoing training and in this case, children with connections to the diabetes care team (family) and medical team play an important role in the management of children’s diabetes. Training and strengthening education helps families to control disease (Sharifirad et al., 2015). Teaching children and their families to improve knowledge and control diabetes and metabolic diseases (Pulungan et al., 2019)

Performing empowerment model lead to increasing mothers’ awareness next, diet treatment of children better control of disease and finally prevent late and early complications of the disease. It should be noted that implementation of this empowerment model is possible with the least equipment also, mothers have more welcome than it and ask for continue that pattern. Therefore, it is suggested such pattern is done to increase the knowledge of mothers and patients’ families in other age groups and other chronically ill in relevant institutes (Deyhoul et al., 2020)

## CONCLUSION

Family centered care for children with diabetes requires family knowledge about care, training skills, building strong motivation for children with diabetes so that complications do not occur. All that must be supported by all family members. Nurses can act as motivators and resource persons in the care of children with diabetes. This is in accordance with the philosophy of pediatric nursing.

## Data Availability

All data produced are available online at
SCOPUS, ProQuest, PubMed, ScienceDirect, SAGEPub, and Google Scholar

https://www.ncbi.nlm.nih.gov/pmc/articles/PMC4389357/

https://www.scopus.com/inward/record.uri?eid=2-s2.0-85041036919&partnerID=40&md5=78857a579588c9339adecebcdae7c474

https://www.frontiersin.org/articles/10.3389/fpsyg.2020.609715/full

https://www.mdpi.com/2227-9067/9/3/445

## ACKNOWLEDGEMENTS

I would like to thank all those who have supported the completion of this article

